# Association between fluoridated toothpaste use and dental caries in Nigeria: A systematic review and meta-analysis

**DOI:** 10.64898/2026.02.27.26346208

**Authors:** Adetayo Aborisade, Amina Mohammed Ali, Mcking Amedari, Abideen Olurotimi Salako, Folahanmi Tomiwa Akinsolu, Olunike Rebecca Abodunrin, Oluwabukola Mary Ola, Mobolaji Timothy Olagunju, George Uchenna Eleje, Joanne Lusher, Oliver Chukwujekwu Ezechi, Moréniké Oluwátóyìn Foláyan

**Author notes:** Correspondence Adetayo Aborisade Department of Oral Diagnostic Sciences Bayero University, Kano, Nigeria.

## Abstract

**Background:** The use of fluoride-containing dentifrices can reduce the risk of dental caries. The systematic review was conducted to address two research questions: (i) the prevalence and frequency of fluoridated toothpaste use among Nigerian children and adolescents across geographic and demographic settings, and (ii) its association with dental caries prevalence, stratified by location and baseline caries risk.

**Methods:** This systematic review, registered with PROSPERO (CRD42022362116), followed the PRISMA guidelines. A PIO framework was applied to include children and adolescents (6 months–19 years) in Nigeria using fluoridated toothpaste, with caries outcomes measured via dmft/DMFT indices. A comprehensive search of PubMed, Web of Science, Scopus, Embase, AJOL, and Google Scholar was conducted from January 2001 to January 2026, supplemented by reference and grey literature searches. Study selection, data extraction, and risk of bias assessment using an adapted Hoy et al. tool were performed independently by multiple reviewers, with high inter-rater reliability (Kappa=0.90). Data were pooled using a random-effects model, with sensitivity, subgroup, and meta-regression analyses conducted to explore heterogeneity and effect modifiers. Publication bias was assessed using funnel plots and Egger’s test.

**Results:** Of 1,194 identified records, 18 studies (n=12,719 participants) were included. The use of fluoridated toothpaste was widespread (prevalence: 61.9% to 95.8%), yet its association with dental caries varied significantly by location. A meta-analysis of 14 studies indicated a significant 16% reduction in caries odds with fluoridated toothpaste use after removal of an influential outlier (OR = 0.84, 95% CI: 0.71–0.99, p=0.04). Subgroup analyses revealed this protective association was significant in urban and rural settings (p<0.05) but absent in suburban Nigeria. Furthermore, dental caries prevalence and severity (DMFT/dmft) were substantially higher in urban and rural areas, where the association was significant, compared to suburban regions. All studies were assessed as having a low risk of bias, and no significant publication bias was detected.

**Conclusion:** Fluoridated toothpaste is widely used in Nigeria and associated with a reduction in the prevalence of dental caries in Nigeria. It appears the relationship is moderated by residential location, and the DMFT/dmft. Longitudinal studies are needed to explore the interactions between the DMFT/dmft, use of fluoridated toothpaste, and residential location in Nigeria.

## Introduction

Dental caries affects about 3.5 billion people globally [1,2]. It is a disease of public health importance because of its link with systemic disorders such as cardiovascular disease, immune system disease, kidney diseases, and mental health [3]; children’s growth and development [4], and poor neurological development [5, 6]. It is also linked with psychological wellness [7,8].

Despite the huge burden of dental caries, it is preventable. One of the most effective tools for preventing dental caries in both primary and permanent dentition is fluoridated toothpaste [9], although its effectiveness increases with higher dmft [10–12]. Fluoride inhibits the metabolic action of cariogenic microbes, such as *Streptococcus mutans*, by inhibiting enzymes involved in acid production [13]. When integrated into the hydroxyapatite crystals of enamel, it generates fluorapatite, which is more resistant to acid assaults [14]. Using fluoridated toothpaste offers a high level of fluoride directly to the tooth surface, promoting remineralization and reducing the risk of dental caries [15].

The widespread use of fluoridated toothpaste has played a significant role in the global reduction of the prevalence of dental caries [11, 16]. However, access to fluoridated toothpaste remains uneven, particularly in low- and middle-income countries such as Nigeria. While the burden of dental caries is relatively low in both the primary [9, 17, 18] and permanent [9, 18] dentitions in Nigeria, the factors contributing to this low burden are poorly understood. The widespread availability of fluoridated toothpaste in Nigeria due to regulations requiring all toothpaste sold in Nigeria to contain fluoride may have contributed to the low prevalence of dental caries, but there is no evidence to support this assertion. In addition, there is a growing interest in non-fluoridated alternatives, driven by concerns about fluoride safety [19, 20]. In addition, the economic downturn and challenges with border control enforcement have increased the availability and purchase of non-fluoridated toothpaste, as non-fluoridated toothpastes are cheaper [21]. This may contribute to a decline in the use of fluoridated toothpaste in Nigeria.

The advocacy for the continued use of fluoridated toothpaste in Nigeria must be evidence-based, although little is known about the specific impact of fluoridated toothpaste on dental caries risk within the Nigerian population. Moreover, there is a lack of context-specific research linking fluoridated toothpaste to dental caries in low- and middle-income countries. Such evidence is crucial for informing evidence-based oral health policy recommendations for low and middle-income countries like Nigeria. Therefore, the objective of this study was to systematically review the existing literature on the association between fluoridated toothpaste use and the prevalence of dental caries in Nigeria.

## Methods

This systematic review was registered with PROSPERO (CRD42022362116). The study was reported following the Preferred Reporting Items for Systematic Reviews (PRISMA) statement and checklist [22]. A 27-item PRISMA checklist is available as an additional file to this protocol in Supplementary File 1.

### Research Questions

Two questions guided the systematic review: (i) What is the prevalence of fluoridated toothpaste use and its frequency among children and adolescents in Nigeria across different geographic and demographic settings? (ii) What is the association between the use of fluoridated toothpaste and the prevalence of dental caries in Nigeria, and how does this relationship vary by geographic location and baseline dental caries risk?

### Population, Intervention, and Outcomes Framework

The Population, Intervention, and Outcomes (PIO) Framework provides a structured approach for specifying the components of this systematic review to determine the prevalence and factors associated with fluoridated toothpaste use and dental caries in Nigeria. (See Table 1).

**Table 1:** PIO Framework.

| Component | Inclusion Criteria |
| --- | --- |
| Participants | Children and adolescents in Nigeria across various settings (rural, urban, and suburban), aged 6 months to 19 years. |
| Intervention | Use of fluoridated toothpaste |
| Outcomes | Prevalence and severity of dental caries, measured by decayed, missing, and filled teeth indices (dmft/DMFT). |

### Search Strategy and Selection of Studies

PubMed, Web of Science (WOS), Embase, EBSCOHost, Scopus, African Journals Online (AJOL), and Google Scholar were searched from January 2001 to January 15, 2026, with the search strategy described in Appendix 1. The search strategy contained terms related to “Prevalence,” “Toothpaste,” “Toothbrushing,” “Fluorides,” “Mouthwashes,” “Oral hygiene,” and “Nigeria.” Alongside electronic database searches, we also reviewed the reference lists of all articles that met the inclusion criteria. This step was taken to identify any potentially relevant studies that might have been missed in the initial search. The included article’s reference list was examined, and studies relevant to our research question were assessed according to the inclusion and exclusion criteria. Additionally, grey literature, including the professional dissertation databases of the National Postgraduate and West African Colleges of Surgeons were also searched for relevant publications. The details of the search strategy are presented in Supplementary File 2.

First, the search outcomes were downloaded to the Rayyan web application (new.rayyan.ai). Next, duplicate items were sorted out and removed. After that, three authors (A.A., A.M.A., and M.A) screened the titles and abstracts of all studies and assessed the full texts of selected studies to determine if they met the inclusion criteria. In addition, information for potentially relevant studies with inaccessible full-text copies was obtained by emailing the authors. The contacted authors responded by providing PDFs or scanned copies of their manuscripts. In cases where multiple publications were associated with the same primary study, a key paper for each primary study was selected, and then the other associated publications were used for supplementary information during the data extraction process. The articles selected independently by the two authors were compared. Disagreements were resolved by discussion or recourse to a fourth review author (M.O.F.). All data items were checked by a fifth reviewer (M.O.F.).

### Eligibility Criteria

All published literature reporting the associations between the use of fluoridated toothpaste and dental caries experience in Nigeria was eligible for study inclusion. Study designs eligible for inclusion were cross-sectional, cohort, and case-control studies. In addition, studies were included if they presented data for the study variables. There was no language restriction.

Studies were excluded if they did not provide information on the sample size, had inaccurate or unavailable outcome data, did not statistically analyse the relationship between sugar consumption and dental caries experience, or featured duplicate samples. Furthermore, review articles, case reports, case series, editorials, laboratory investigations, or reviews devoid of primary data were excluded. The details of the excluded studies and reasons are presented in Supplementary File 3.

### Data Extraction

Two independent reviewers (A.A and A.M.A) used a pretested data extraction form prepared in Microsoft Excel to extract details of articles that met the inclusion criteria independently. The information extracted was the author’s name, year of article publication, study design, study location, and study setting. Information on the study participants (sample size, age range, and sex distribution), the prevalence of dental caries, the DMFT/dmft, and their frequency of use of fluoridated toothpaste were also extracted. Discrepancies among reviewers during the extraction process were resolved by a third reviewer (M.A). The details of the extraction are available in Supplementary File 4.

### Risk of Bias Assessment

An adapted version of the risk of bias tool for prevalence studies, developed by Hoy and colleagues [23] was used to assess the quality and risk of bias for studies that met the inclusion criteria. Each study underwent scrutiny across nine domains of bias, namely: description of the target population of the study, the sampling frame, sampling techniques, response rate, non-proxy collection of data, case definition of the study, validity, and reliability of the study instrument, mode of data collection, and appropriate description of numerator and denominator for the parameter of interest. The total score ranged from 0 to 9, with the overall score categorized as follows: 0–3: “low risk,” 4–6: “moderate risk,” and 7–9: “high risk” of bias. The assessment was done by three independent reviewers (A.A., A.M.A., and M.A), and discrepancies were resolved by a fourth reviewer (M.O.F).

### Inter-rater Reliability

Inter-rater reliability between the two reviewers (A.A. and A.M.A.) was 91%, with a Kappa score of 0.90 for screening titles and abstracts and a Kappa score of 0.90 for the included full-text articles. Kappa’s excellent reliability score ranges from 0.81 to 1.0 [24].

### Statistical Analysis

Relevant studies were synthesised using a random-effects model to account for heterogeneity. The independent variable was the use of fluoridated toothpaste. The dependent variable was caries prevalence. Odds ratios (ORs) with 95% confidence intervals served as the summary measure. These were calculated from extracted frequencies and denominators reported in the selected studies.

The pooled estimate of the summary measure was generated using the Mantel-Haenszel method. Only studies reporting binary data on caries presence or absence were included. Study heterogeneity was evaluated using Cochran’s Q.

The Q test, the I² statistic, and visual inspection of forest plots were used for assessing heterogeneity. A p-value less than 0.05 for the Q test was considered statistically significant. I² values above 80% indicated substantial heterogeneity. All analyses were conducted using R and RStudio Version 4.5.1 (Great Square Root) with the Meta, Metafor, and Dmetar packages.

### Sensitivity Analysis and Outlier Analysis

Studentized residuals for study parameters were computed. Studies with absolute residuals exceeding 2 were considered outliers. A leave-one-out sensitivity analysis was then performed to identify influential studies. Identified influential studies were removed, and summary measures were recalculated. These studies were also excluded from all subsequent analyses to minimise potential sources of heterogeneity.

### Moderator Analysis

When substantial heterogeneity was identified, subgroup analyses were conducted to determine its sources. Subgroup analyses were performed by geopolitical zone and by study design. Study design categories included population-based, hospital-based, or other (including school-based) studies. Additional subgroup analyses were conducted by dentition type: primary, mixed, or permanent. To further explore heterogeneity, univariate meta-regression was performed by year of publication and study sample size. This was done to evaluate their significance in the pooled summary estimate and the association between fluoridated toothpaste use and caries prevalence.

### Publication Bias

Publication bias in this study was assessed visually using funnel plots. It was also assessed objectively using rank correlation and Egger’s regression test. The significance of both objective tests was set at p<0.05.

## Results

### Selection of Studies

As shown in Figure 1, 1194 records were retrieved from both the databases and the grey literature resources. After removing duplicates, 946 records remained eligible for title and abstract screening. Of these, 893 studies were excluded, and the full-text records of 56 studies were reviewed. Eighteen studies met the eligibility criteria [17, 25–41].

**Figure 1:**
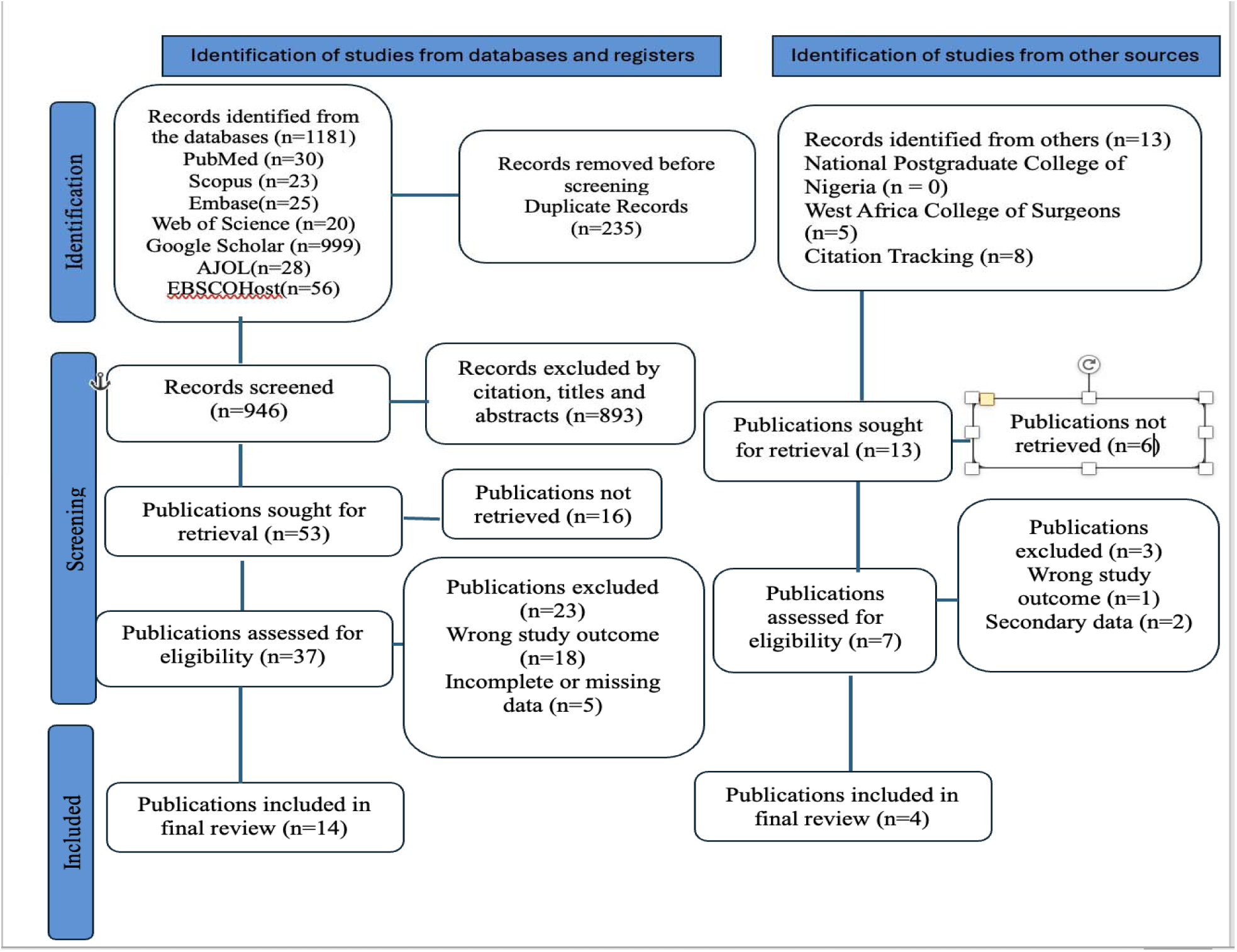
PRISMA Flowchart of the Publication Selection Process.

### Characteristics of Included Studies

Table 2 presents the extracted data from the 18 studies. The studies were conducted between 2011 and 2025. Five studies (27.8%) were conducted between 2011 and 2019 [17, 25–28], six studies (33.3%) between 2020 and 2021 [29–34], and the remaining seven studies (38.9%) between 2022 and 2025 [35–41]. These findings indicate a consistent increase in publications addressing the impact of fluoridated toothpaste on dental caries prevalence.

**Table 2:**
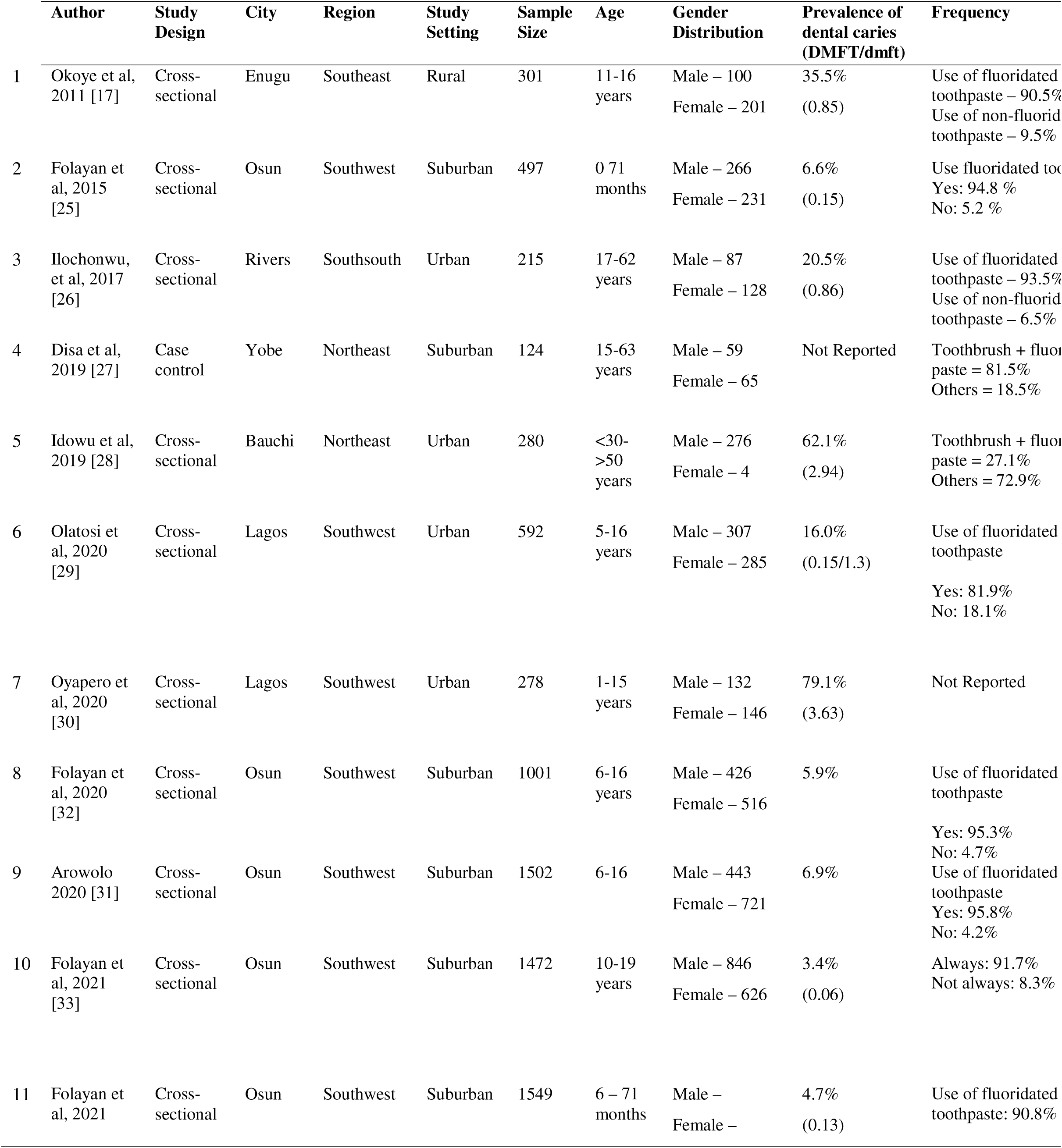
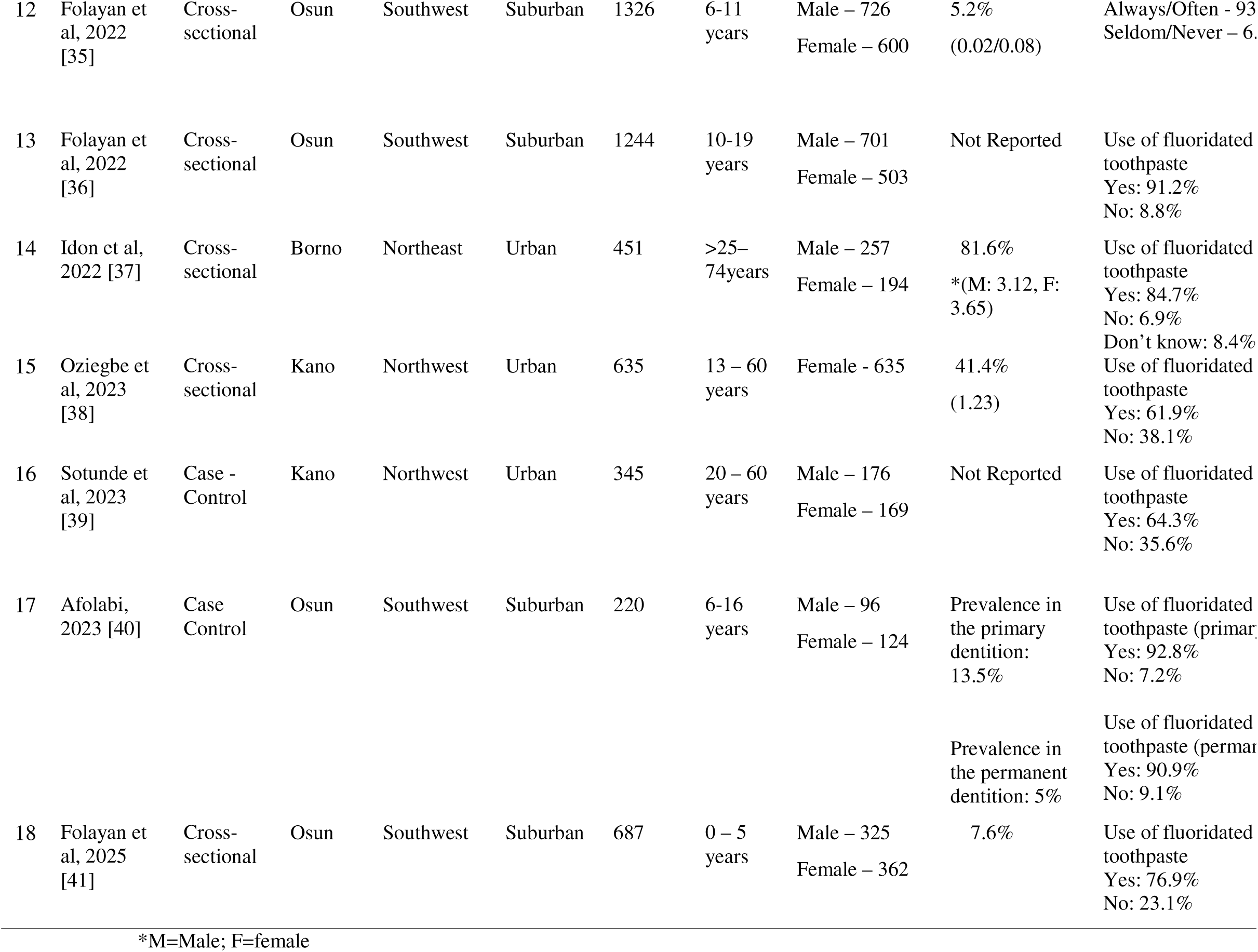
Characteristics of Included Studies.

|  | Author | Study Design | City | Region | Study Setting | Sample Size | Age | Gender Distribution | Prevalence of dental caries (DMFT/dmft) | Frequency |
| --- | --- | --- | --- | --- | --- | --- | --- | --- | --- | --- |
| 1 | Okoye et al, 2011 [17] | Cross-sectional | Enugu | Southeast | Rural | 301 | 11-16 years | Male – 100<br>Female – 201 | 35.5%<br>(0.85) | Use of fluoridated toothpaste – 90.5%<br>Use of non-fluorid toothpaste – 9.5% |
| 2 | Folayan et al, 2015 [25] | Cross-sectional | Osun | Southwest | Suburban | 497 | 0-71 months | Male – 266<br>Female – 231 | 6.6%<br>(0.15) | Use fluoridated toothpaste: Yes: 94.8 %<br>No: 5.2 % |
| 3 | Ilochonwu, et al, 2017 [26] | Cross-sectional | Rivers | Southsouth | Urban | 215 | 17-62 years | Male – 87<br>Female – 128 | 20.5%<br>(0.86) | Use of fluoridated toothpaste – 93.5%<br>Use of non-fluorid toothpaste – 6.5% |
| 4 | Disa et al, 2019 [27] | Case control | Yobe | Northeast | Suburban | 124 | 15-63 years | Male – 59<br>Female – 65 | Not Reported | Toothbrush + fluoridated toothpaste = 81.5%<br>Others = 18.5% |
| 5 | Idowu et al, 2019 [28] | Cross-sectional | Bauchi | Northeast | Urban | 280 | <30-<br>>50 years | Male – 276<br>Female – 4 | 62.1%<br>(2.94) | Toothbrush + fluoridated toothpaste = 27.1%<br>Others = 72.9% |
| 6 | Olatosi et al, 2020 [29] | Cross-sectional | Lagos | Southwest | Urban | 592 | 5-16 years | Male – 307<br>Female – 285 | 16.0%<br>(0.15/1.3) | Use of fluoridated toothpaste<br>Yes: 81.9%<br>No: 18.1% |
| 7 | Oyapero et al, 2020 [30] | Cross-sectional | Lagos | Southwest | Urban | 278 | 1-15 years | Male – 132<br>Female – 146 | 79.1%<br>(3.63) | Not Reported |
| 8 | Folayan et al, 2020 [32] | Cross-sectional | Osun | Southwest | Suburban | 1001 | 6-16 years | Male – 426<br>Female – 516 | 5.9% | Use of fluoridated toothpaste<br>Yes: 95.3%<br>No: 4.7% |
| 9 | Arowolo 2020 [31] | Cross-sectional | Osun | Southwest | Suburban | 1502 | 6-16 | Male – 443<br>Female – 721 | 6.9% | Use of fluoridated toothpaste<br>Yes: 95.8%<br>No: 4.2% |
| 10 | Folayan et al, 2021 [33] | Cross-sectional | Osun | Southwest | Suburban | 1472 | 10-19 years | Male – 846<br>Female – 626 | 3.4%<br>(0.06) | Always: 91.7%<br>Not always: 8.3% |
| 11 | Folayan et al, 2021 | Cross-sectional | Osun | Southwest | Suburban | 1549 | 6 – 71 months | Male –<br>Female – | 4.7%<br>(0.13) | Use of fluoridated toothpaste: 90.8% |
| 12 | Folayan et al, 2022 [35] | Cross-sectional | Osun | Southwest | Suburban | 1326 | 6-11 years | Male – 726<br>Female – 600 | 5.2%<br>(0.02/0.08) | Always/Often - 93<br>Seldom/Never – 6. |
| 13 | Folayan et al, 2022 [36] | Cross-sectional | Osun | Southwest | Suburban | 1244 | 10-19 years | Male – 701<br>Female – 503 | Not Reported | Use of fluoridated toothpaste<br>Yes: 91.2%<br>No: 8.8% |
| 14 | Idon et al, 2022 [37] | Cross-sectional | Borno | Northeast | Urban | 451 | >25–74years | Male – 257<br>Female – 194 | 81.6%<br>*(M: 3.12, F: 3.65) | Use of fluoridated toothpaste<br>Yes: 84.7%<br>No: 6.9%<br>Don't know: 8.4% |
| 15 | Oziegbe et al, 2023 [38] | Cross-sectional | Kano | Northwest | Urban | 635 | 13 – 60 years | Female - 635 | 41.4%<br>(1.23) | Use of fluoridated toothpaste<br>Yes: 61.9%<br>No: 38.1% |
| 16 | Sotunde et al, 2023 [39] | Case - Control | Kano | Northwest | Urban | 345 | 20 – 60 years | Male – 176<br>Female – 169 | Not Reported | Use of fluoridated toothpaste<br>Yes: 64.3%<br>No: 35.6% |
| 17 | Afolabi, 2023 [40] | Case Control | Osun | Southwest | Suburban | 220 | 6-16 years | Male – 96<br>Female – 124 | Prevalence in the primary dentition: 13.5%<br>Prevalence in the permanent dentition: 5% | Use of fluoridated toothpaste (primary)<br>Yes: 92.8%<br>No: 7.2%<br>Use of fluoridated toothpaste (permanent)<br>Yes: 90.9%<br>No: 9.1% |
| 18 | Folayan et al, 2025 [41] | Cross-sectional | Osun | Southwest | Suburban | 687 | 0 – 5 years | Male – 325<br>Female – 362 | 7.6% | Use of fluoridated toothpaste<br>Yes: 76.9%<br>No: 23.1% |

\*M=Male; F=female

Sample sizes in the included studies ranged from 124 to 1,549, with a total of 12,719 participants (5,223 males and 5,316 females). One study [36] did not report the male-female ratio, while another [38] focused exclusively on a female population. Participants’ ages ranged from 6 months to 74 years. Nine studies (50%) primarily examined permanent teeth [17, 26–28, 33, 36–39], and four studies (22.2%) addressed both the primary [25, 31, 34, 41] and mixed dentition [29, 30, 32, 35] respectively. One study [40] reported on both the primary and permanent dentition.

Among the 18 studies analyzed, 11 (61.1%) were conducted in the Southwest geopolitical zone of Nigeria, with nine in Osun State [25, 31–36, 40, 41] and two in Lagos State [29, 30]. Three studies were conducted in the Northeast zone, specifically in Yobe [27], Borno [37], and Bauchi [28] states. Two studies were conducted in Kano state [38, 39], and one study was conducted in Enugu [17] and Rivers [26], each representing the Northwest, Southeast, and South-south geopolitical zones, respectively.

The majority of studies were conducted in suburban (n=10, 55.6%) and urban (n=7, 38.9%) settings, with only one study in a rural area [17]. Nine (50%) of the studies were community-based [17, 25, 33–36, 38, 39, 41], five (27.8%) were hospital-based [26, 27, 30, 37, 40], and three (16.7%) were school-based [29, 31, 32]. All included studies demonstrated a low risk of bias, as detailed in Table 2.

### Prevalence of Dental Caries

Table 2 indicates that dental caries prevalence ranged from 3.4% [33] to 81.6% [37]. Among preschool children aged 6 to 71 months, prevalence ranged from 4.7% [34] to 13.5% [40]. In mixed dentition, prevalence was 5.2% [35] to 79.1% [30], while in permanent dentition, it ranged from 3.4% [33] to 81.6% [37]. In rural Nigeria, dental caries prevalence was 35.5% [17] among children in secondary schools. In urban areas, prevalence ranged from 16.0% among children in private primary schools [29] to 81.6% in the hospital-based study [37]. In suburban areas, prevalence varied from 3.4% [33] to 13.5% [40]. Overall, hospital-based studies [26, 27, 30, 37, 40] reported a higher caries prevalence compared to the school [29, 31, 32] and community-based studies [17, 25, 33–36, 38, 39, 41].

### Prevalence of Use of Fluoridated Toothpaste

Table 2 shows that the prevalence of fluoridated toothpaste use in Nigeria ranged from 61.9%[38] to 95.8% [31]. In suburban areas, usage rates were between 76.9% [41] and 95.8% [31]. The only exception was the study conducted in an institutional setting, which reported a lower usage rate of 27.1% [28]. In the single rural study, prevalence was 90.5% [17], while in the urban areas, it ranged from 61.9% [38] to 93.5% [26].

Among preschool children aged 6 to 71 months, the prevalence of fluoridated toothpaste use ranged from 76.9% [41] to 95.8% [31]. For children with mixed dentition, usage ranged from 81.9% [29] to 95.3% [32]. For the permanent dentition, prevalence ranged from 61.9% [38] to 93.5% [26], excluding the study conducted in an institutional setting [28].

### The DMFT/dmft profile

The dmft index ranged from 0.06 in suburban Nigeria [33] to 1.23 [38] in urban Nigeria, while the DMFT index ranged from 0.02 in suburban areas [35] to 3.63 in urban areas [37]. In studies reporting both indices [29, 35], dmft values were higher than DMFT values. While no study explicitly analyzed the relationship between fluoridated toothpaste use and DMFT/dmft indices, the data suggest a contextual trend. Elevated DMFT/dmft values corresponded with geographic settings (urban and rural) in which fluoridated toothpaste use was significantly associated with lower caries prevalence. Conversely, in suburban regions where no significant association was found, DMFT/dmft scores were generally lower.

### Fluoridated Toothpaste Use and Its Association with Dental Caries

Table 2 presents findings from five studies among preschool children (6-71 months) in suburban Nigeria [25, 31, 34, 40, 41], indicating no statistically significant association between fluoridated toothpaste use and dental caries. Although the odds ratios suggested a reduced risk of dental caries with fluoridated toothpaste, these results did not reach statistical significance.

Among children with mixed dentition, three of four studies [30, 32, 35] found no significant association between fluoridated toothpaste use and dental caries experience. In contrast, one study [29] reported a significantly higher prevalence of caries among users of non-fluoridated toothpaste. These studies were conducted in both suburban and urban regions of Nigeria.

In the permanent dentition, results differed by geographic location. In urban Nigeria, one of five studies [29] identified a significant association between fluoridated toothpaste use and dental caries experience. No significant associations were reported in the four studies conducted in suburban Nigeria [27, 33, 36, 40]. Conversely, in rural Nigeria [17], children who used fluoridated toothpaste had statistically significantly lower odds of dental caries experience.

### Quality Assessment

Supplemental File 1 and Table 3 provide a summary of the assessment outcomes. All included studies were determined to have an assessment outcome. All included studies were determined to have a low risk of bias.

**Table 3:**
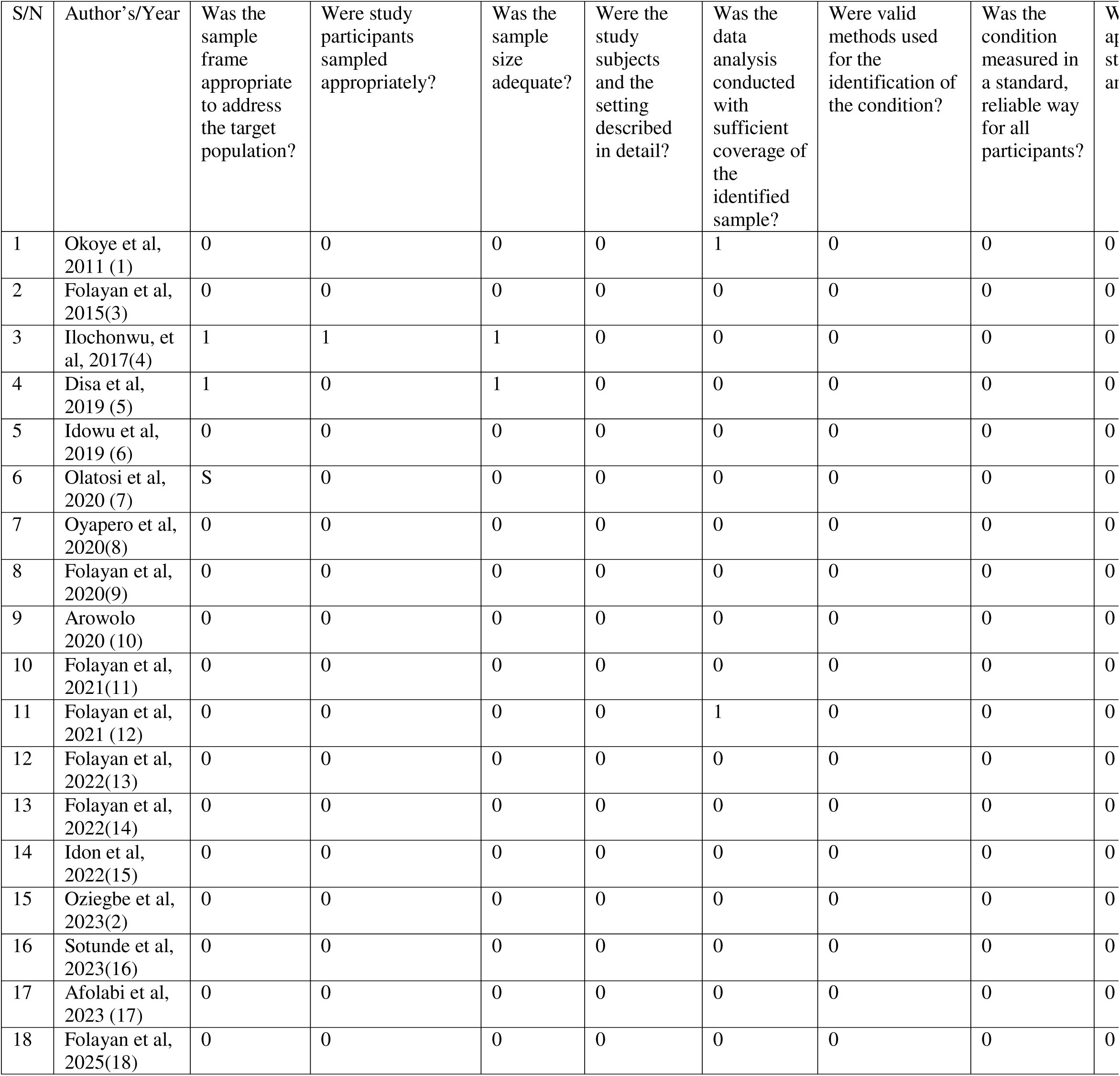
Quality Assessment.

### Meta-analysis of the association between the use of fluoridated toothpaste and dental caries

Fourteen studies reporting on the association between fluoridated toothpaste use and dental caries were pooled in a random-effects meta-analysis [17, 25, 27–29, 31–33, 35–37, 39–41]. Overall, the odds of dental caries were 31% lower among participants who used fluoridated toothpaste than among those who did not (OR = 0.69, 95% CI: 0.43–1.12; p = 0.123). Heterogeneity across studies was very high (I² = 81.7% 95% CI: 70.8–88.5; p < 0.0001). This is shown in Figure 2

**Figure 2:**
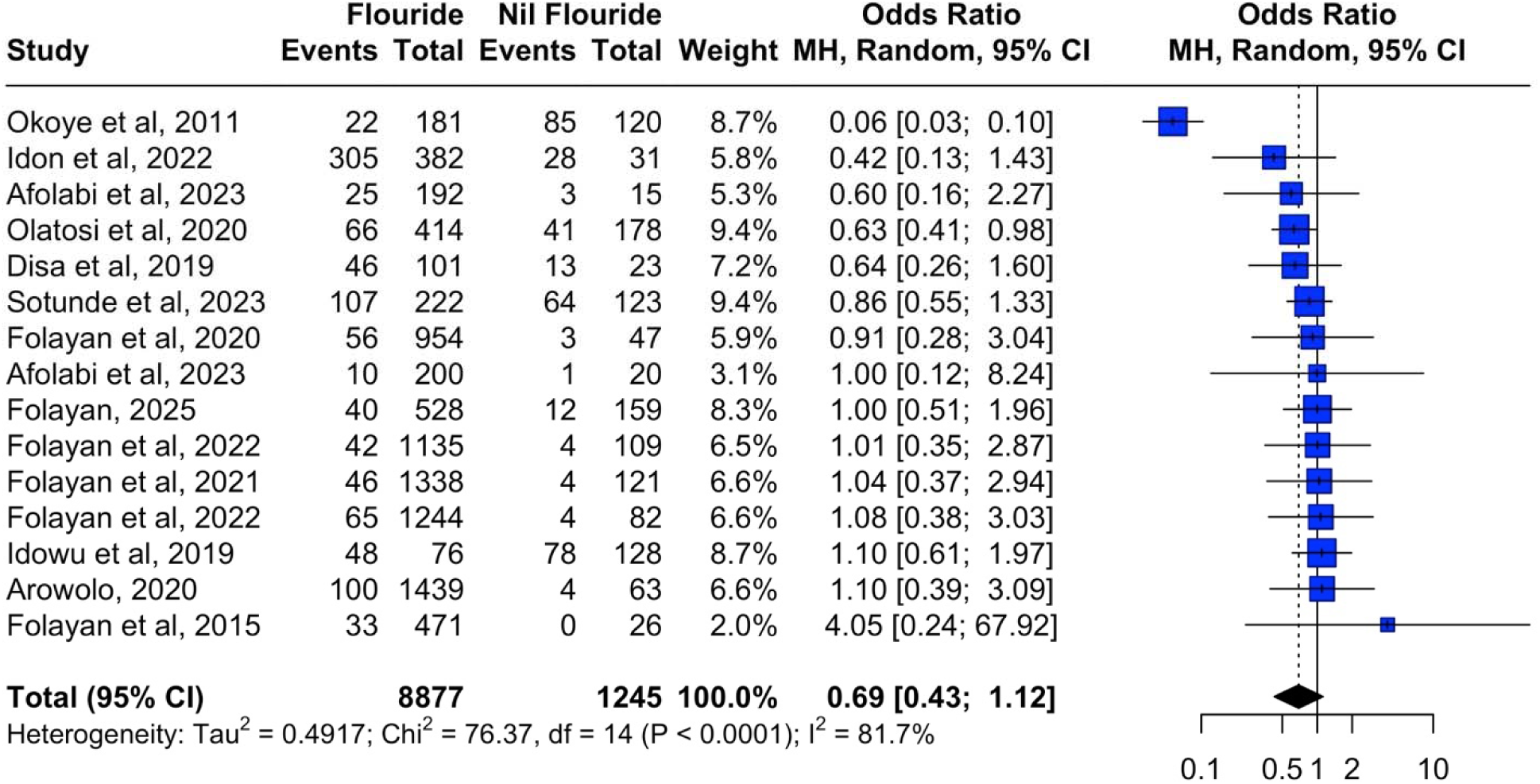
Forest plot of fluoridated toothpaste use and dental caries.

Computation of studentized residuals and the leave-one-out sensitivity analysis revealed one study to be both an outlier and an influential study [17], as seen in the Baujat plot (Figure 3) and the Influential plot (Figure 4). The study was removed, and the pooled OR was re-computed. The recomputed pooled OR showed that the odds of having caries were 16% lower for individuals who use fluoridated toothpaste (0.84; 95% CI: 0.71 - 0.99; p=0.04). The 1^2^ was 0.0% to indicate the consistency across the remaining studies in this meta-analysis.

**Figure 3:**
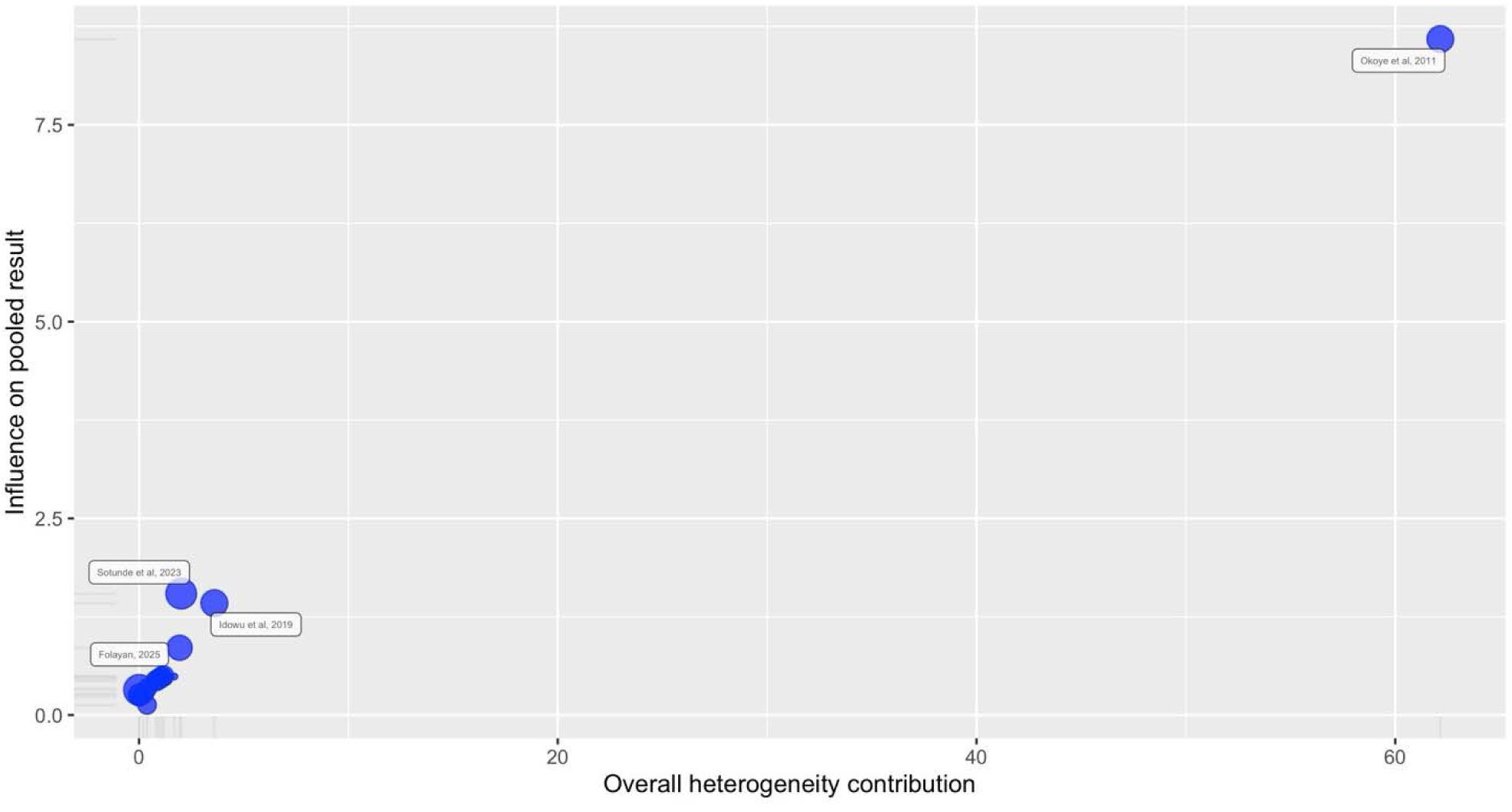
The Baujat plot, revealing the influential study.

**Figure 4:**
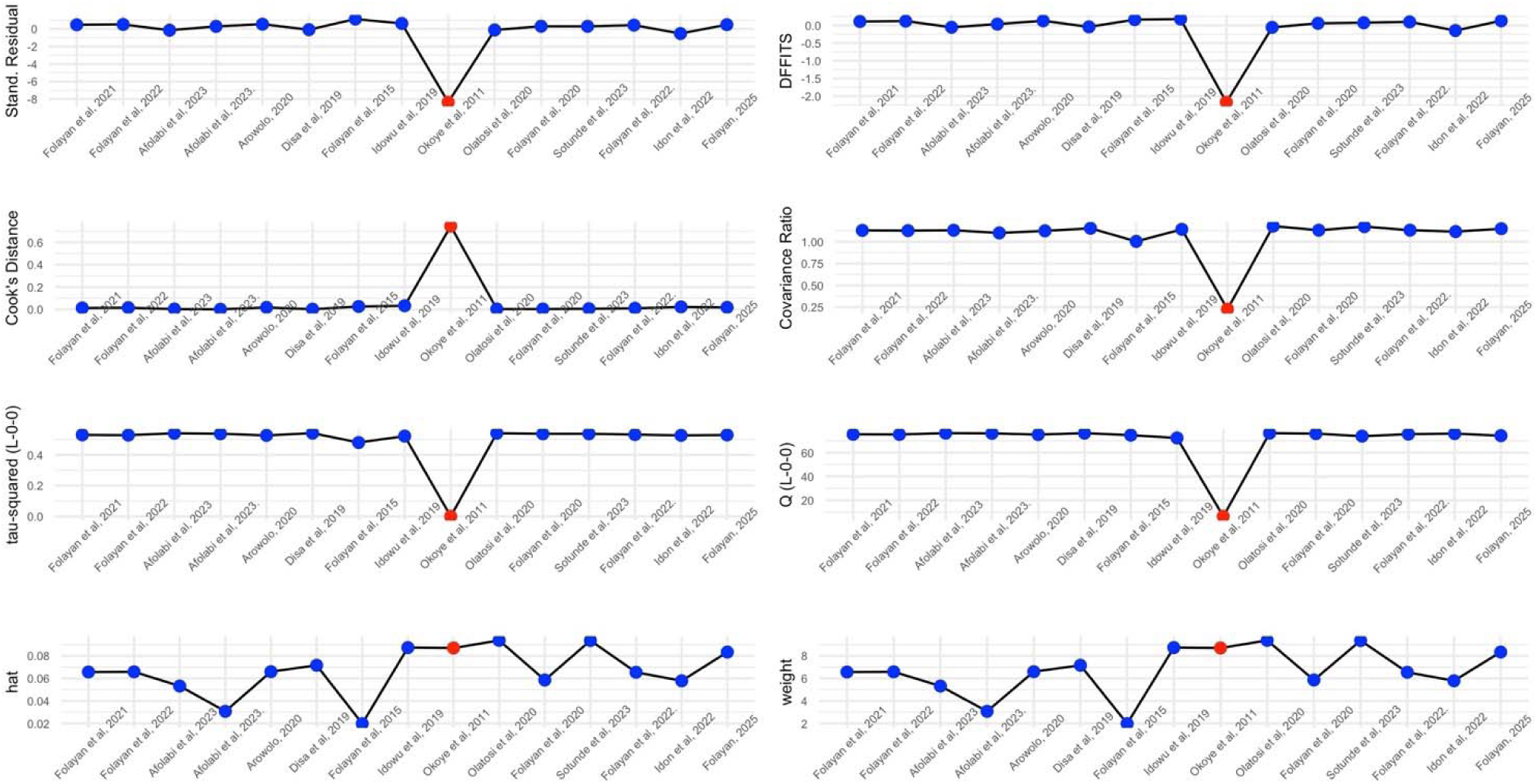
The influential plot showing the outlier study (in red)

### Subgroup Analysis

Subgroup analysis performed across the types of dentitions showed that the odds of dental caries were 26% lower among participants who used fluoridated toothpaste in the mixed dentition, while the odds were lower at 13% in the permanent dentition. There was no difference in the odds ratio in caries prevalence in the primary dentition. The difference across the types of dentitions was not statistically significant (Figure 5).

**Figure 5:**
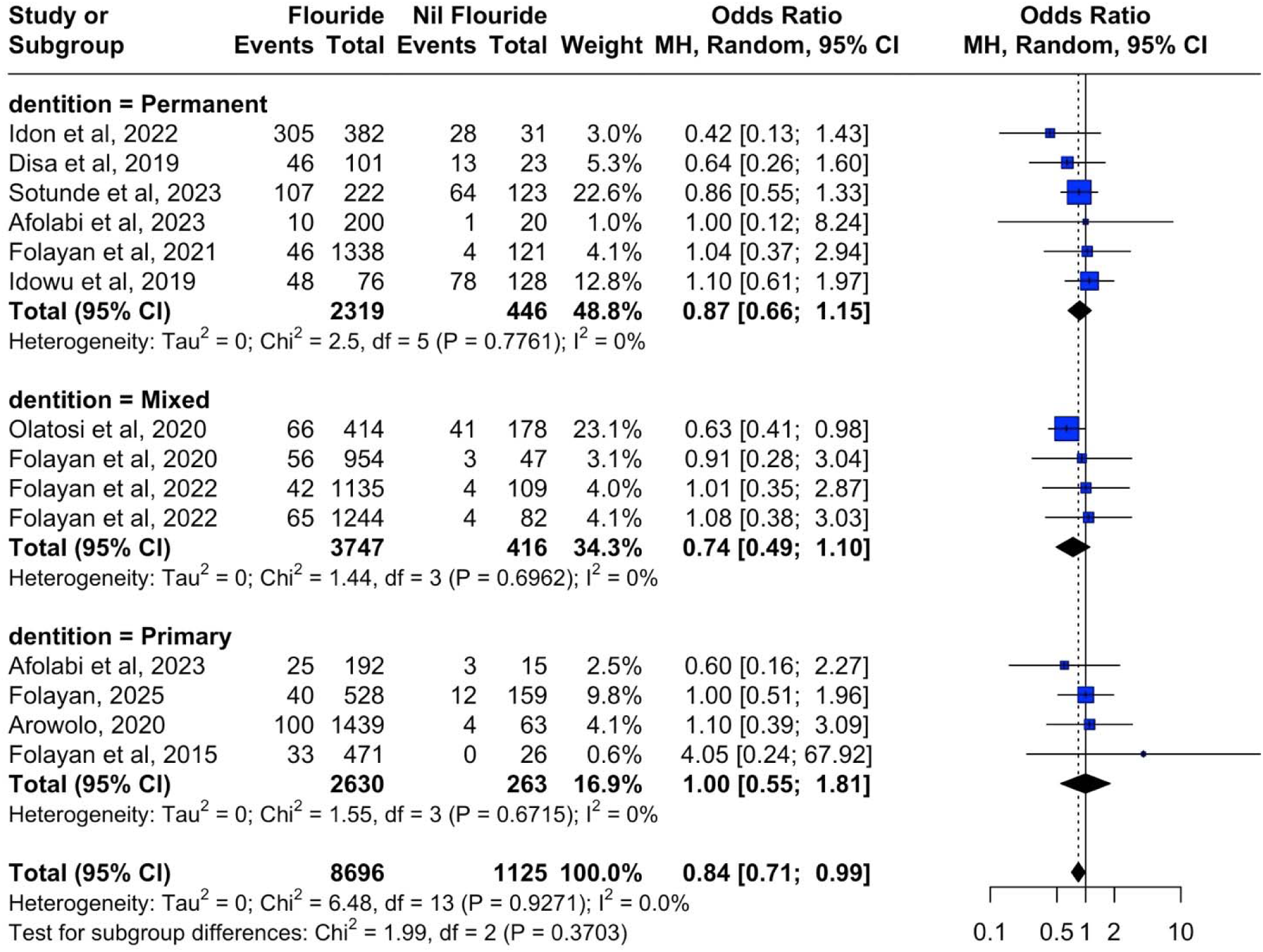
Forest plot showing the subgroup analysis across type of dentitions.

There was a statistical difference, however, across the study designs. Hospital-based studies reported 41% lower odds of caries prevalence in individuals who used fluoridated toothpaste, while population-based designs showed 5% lower odds for dental caries experience. This difference across study designs was statistically significant (p=0.008) as shown in Figure 6.

**Figure 6:**
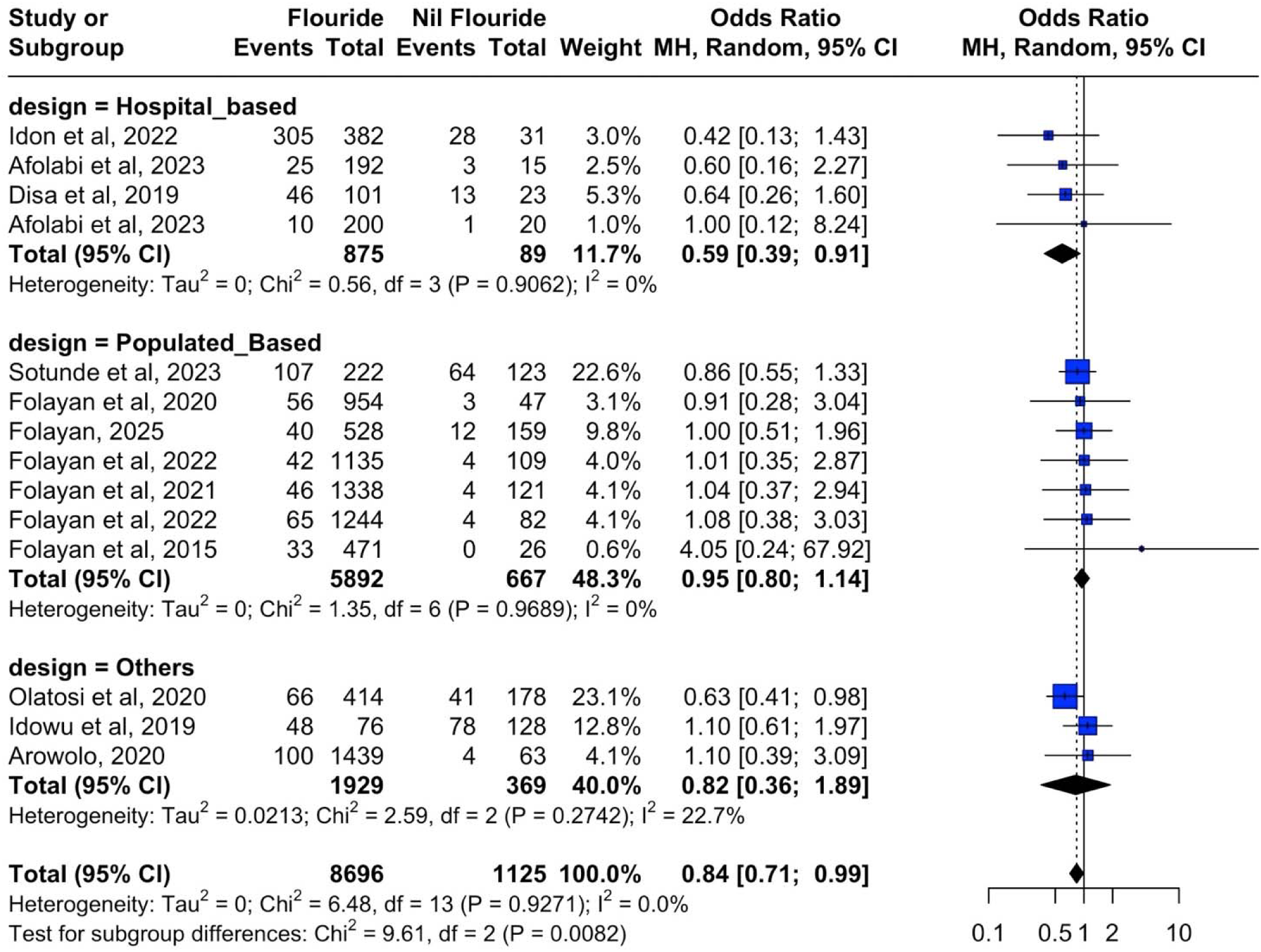
Forest plot showing the subgroup analysis across study designs.

Univariate meta-regression using year of publication (p=0.89) and study sample size (p=0.29) showed that neither variable had a significant moderating effect on the association between use of fluoridated toothpaste and caries prevalence.

Analysis also showed that the odds of dental caries were consistent across the Southern and Northern regions of Nigeria, with 17% and 15% lower odds of caries prevalence in both regions, respectively, with the use of fluoridated toothpaste (Figure 7).

**Figure 7:**
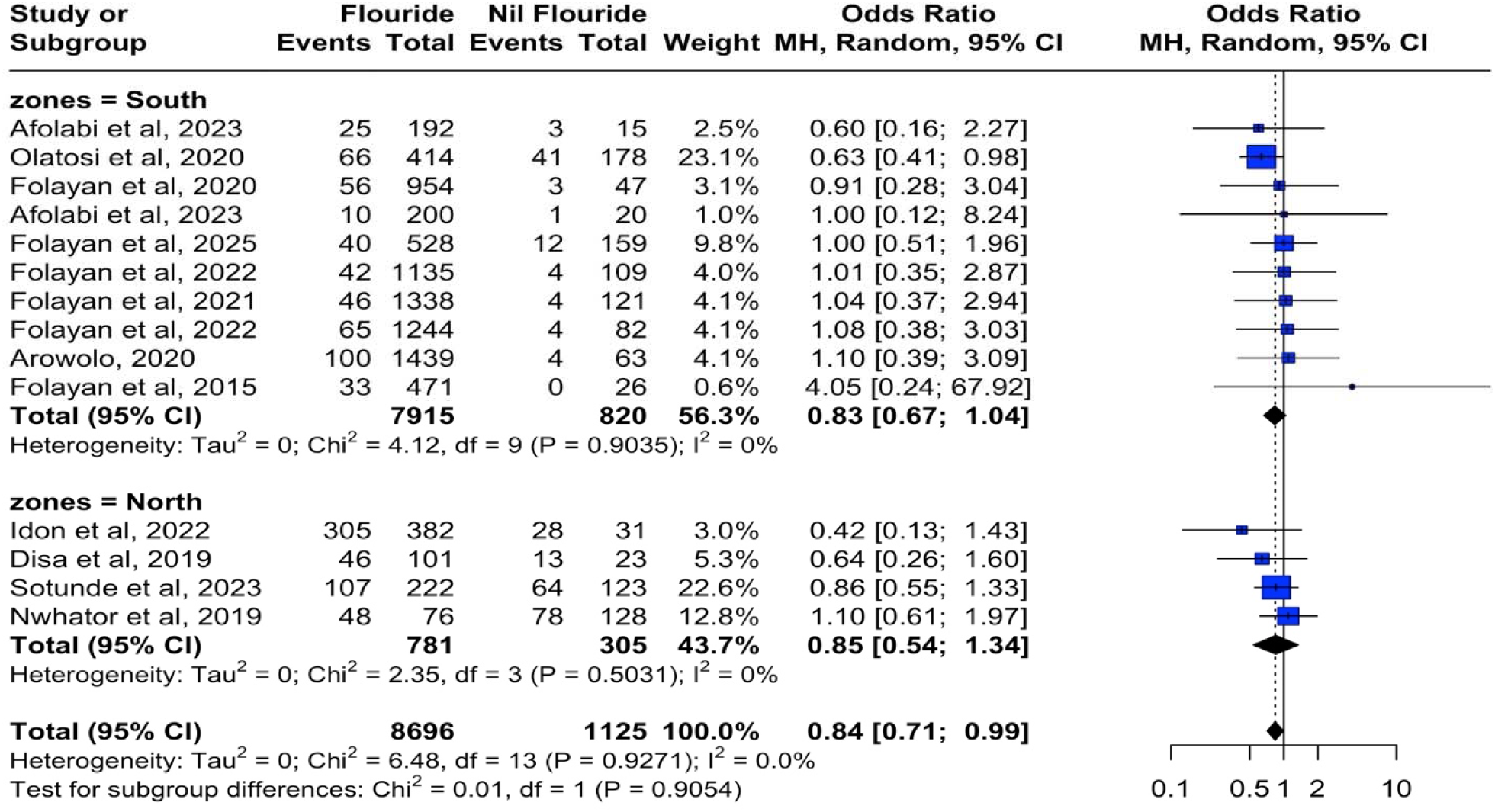
Forest plot showing the subgroup analysis across regions in Nigeria Publication Bias.

A funnel plot of the included studies did not reveal clear evidence of asymmetry (Figure 8). Visual inspection suggested that the studies were distributed approximately symmetrically around the pooled odds ratio. Both the rank correlation test (p=0.87) and Egger’s regression test (p=0.32) were not significant.

**Figure 8:**
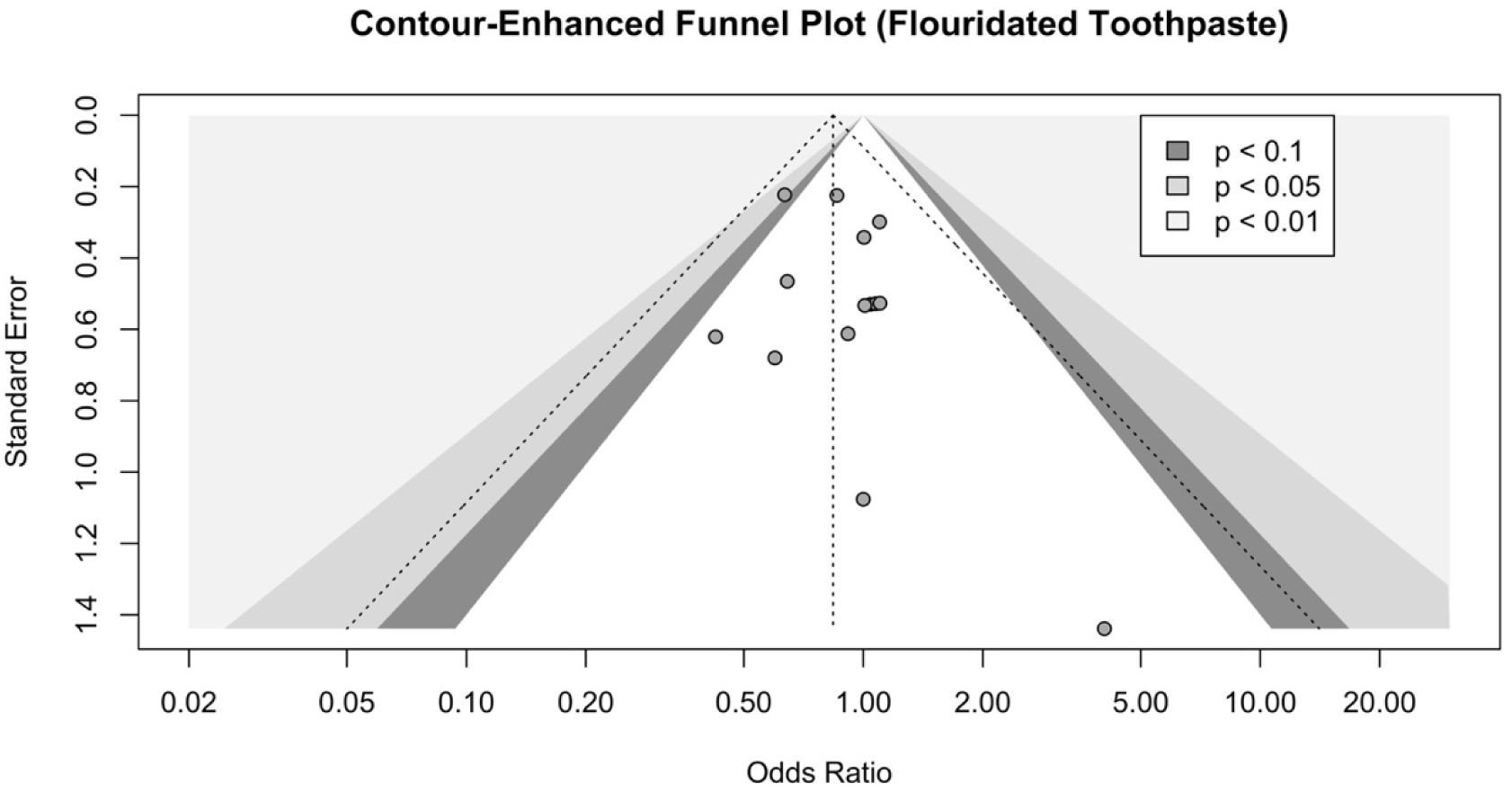
Funnel plot of publication bias for fluoridated toothpaste studies.

## Discussion

The systematic review findings indicate that fluoridated toothpaste use is widespread and is highest in semi-urban and urban areas. However, its association with dental caries was inconsistent: while significant protective effects were observed in rural and urban settings, no significant association was found in semi-urban regions. The results suggest that the relationship between fluoridated toothpaste use and caries prevalence may be moderated by residential location and baseline caries risk, with stronger effects observed in higher-risk environments.

Strengths of this review include a comprehensive search, rigorous risk of bias assessment, and subgroup analyses exploring heterogeneity. In addition, the study included a range of age groups, from young children to adolescents, providing a broad perspective on fluoride use and dental caries across different developmental stages. This allows for a more comprehensive understanding of how fluoride use habits develop and change with age and how these habits impact dental health. In addition, the study’s use of observational data from studies with a low risk of bias enhances the reliability of the findings [42–45].

Limitations include the predominance of cross-sectional designs, regional overrepresentation (particularly Southwest Nigeria), and a lack of adjustment for key confounders such as diet and socioeconomic status [46, 47], which restricts causal inference and generalizability [48]. The cross-sectional nature of the studies also prevents assessing how fluoride use and dental caries prevalence change over time [49]. In addition, the studies were predominantly conducted in suburban settings in Southwest Nigeria, and two of the six geopolitical zones in Nigeria were unrepresented in the data. Furthermore, the data largely pertained to infants, toddlers, preschoolers, school-age children, and adolescents, with no information available for adults. Despite these limitations, the study provides new data on the link between the use of fluoridated toothpaste and dental caries in Nigeria.

The findings from this study align with those of other systematic reviews that showed a protective effect of fluoridated toothpaste against dental caries, often reporting significant reductions in dental caries incidence with regular fluoride use [50–54]. Daily fluoride use in various forms (toothpaste, mouth rinses, supplements) is a critical component of dental caries prevention [55]. However, the findings from the current study align more closely with reviews from other low- and middle-income countries, where the protective effect of fluoride is less consistent [56, 57]. These discrepancies can be attributed to several factors, including frequency of use, oral hygiene practices, and access to dental care services [58]. In many low- and middle-income countries, additional factors such as poor nutrition, high sugar consumption, and limited access to professional dental care can undermine the benefits of fluoride, leading to less significant outcomes in dental caries prevention [59, 60].

This systematic review elucidates the nuanced relationship between fluoridated toothpaste use and dental caries prevalence among children and adolescents in Nigeria. While fluoridated toothpaste use is widespread, its association with caries prevention is not uniform; it appears significantly protective in rural and urban settings but not in suburban areas. To interpret these context-dependent findings, we employ two complementary theoretical frameworks: the Socio-Ecological Model (SEM) [61] and the Diffusion of Innovations (DoI) Theory [62].

The SEM provides a multi-level lens to understand how individual behaviors interact with broader environmental, social, and policy factors. At the individual and interpersonal level, the high prevalence of fluoride use in suburban areas coexists with low caries rates but without a statistically significant association, suggesting possible saturation effects [63] and the influence of superior oral health literacy and dietary habits that may dilute the measurable impact of fluoride alone [64].

At the community and institutional level, the suburban settings where the studies were conducted often benefit from enhanced access to dental facilities and school-based preventive programs [65, 66], which may contribute to uniformly low caries rates irrespective of fluoride use. These studies were conducted in a small university town with two primary health centers offering dental services, a tertiary dental hospital, and a dental school, which likely increased the population’s access to oral health information, preventive care, and treatment. These resources have the potential to influence the findings. Therefore, future research might compare these results with those of other suburban settings in Nigeria.

In contrast, rural and urban areas, which exhibited higher baseline caries, showed a stronger protective effect from fluoridated toothpaste, indicating that community infrastructure, including healthcare access and the retail environment, may modulate fluoride’s preventive role [67]. At the policy level, although national regulations mandate fluoride in toothpaste, variability in enforcement, affordability, and the presence of informal markets likely influence consistent usage, particularly in resource-constrained households, making fluoride a more salient factor in higher-risk settings [68].

This does not necessarily imply that the use of fluoride-containing dentifrices is not important in reducing the risk of dental caries in suburban populations in Nigeria. The study findings, however, suggest that in resource-constrained settings where strategic allocation of resources is critical, prioritizing access to fluoridated toothpaste for rural and urban communities over suburban ones may be more effective for dental caries control. The DoI theory offers further insight into the adoption and impact of fluoridated toothpaste. In suburban Nigeria, fluoride toothpaste has reached the late majority stage of adoption, with near-universal use potentially reducing its relative advantage and observable isolated effect. In higher-risk rural and urban settings, the perceived benefit is likely greater, reinforcing consistent use and yielding more detectable outcomes [69, 70]. Our meta-analysis indicated an overall protective effect, but this was heavily influenced by contextual moderators, underscoring that the realized benefits of fluoride are substantially shaped by local socioecological conditions.

These findings should be interpreted cautiously due to the complex interplay of etiological and risk factors not fully addressed in the reviewed observational studies. The variability in these findings underscores the imperative for context-sensitive oral health strategies. Rather than a one-size-fits-all promotion of fluoridated toothpaste, public health approaches should be tailored. In high-caries rural and urban communities, efforts should strengthen access through subsidy programs and integrate distribution into maternal-child health initiatives. In lower-risk suburban areas, the focus could shift toward multifactorial prevention, including sugar reduction and fissure sealants, while maintaining fluoride use as a foundational element [47, 71]. Further research is needed to assess the generalizability of these results and identify the factors influencing the relationship between fluoridated toothpaste use and dental caries in Nigeria.

Significant knowledge gaps remain, however, particularly regarding adult populations and the northern geopolitical zones of Nigeria. The socioeconomic and cultural heterogeneity across the country necessitates caution in generalizing these findings and highlights the need for research that includes diverse regions and age groups [72]. Future studies should employ longitudinal and quasi-experimental designs to establish causality and better explore mediators such as socioeconomic status and dietary patterns. Investigating community-level fluoridation policies and the cultural determinants of fluoride adherence through mixed-methods approaches is also warranted [73, 74].

In conclusion, this review advances the understanding of fluoride toothpaste efficacy within the complex socioecological landscape of a low- and middle-income country. By applying the SEM and DoI theory, we move beyond simple binary associations toward a nuanced, systems-oriented perspective. For Nigeria and similar settings, oral health strategies must be theoretically informed, context-aware, and tailored to local determinants to effectively reduce disparities and maximize preventive impact.

## Author Contributions

Conceptualization, M.O.F.; methodology, A.A., A.M.A. & M.A..; software, A.A., A.M.A. & M.A.; validation, F.T.A., M.O.F. and G.U.E.; formal analysis, A.A., A.M.A. & M.A..; investigation, F.T.A., O.R.A. & O.M.O.; resources, M.T.O.; data curation, M.T.O. & O.M.O.; writing—original draft preparation, A.A., A.M.A, M.A. & M.O.F.; writing—review and editing, F.T.A., O.R.A., O.M.O., J.L., O.C.E. & M.O.F.; visualization, M.O.F.; supervision, M.O.F.; project administration, M.O.F.; funding acquisition, M.O.F.. All authors have read and agreed to the published version of the manuscript.

## Acknowledgments

Not applicable.

## Data Availability Statement

The datasets analyzed for this study are publicly accessible.

