## Supplemental Files 1 - 4 for "Association between fluoridated toothpaste use and dental caries in Nigeria: A systematic review and meta-analysis": Supplemental File 3.docx

**Supplemental File 2: Summary of Excluded Studies**

| S/N | Title | Year | Author | Reasons for Exclusion |
| --- | --- | --- | --- | --- |
| 1 | Dental caries occurrence and associated oral hygiene practices among rural and urban Nigerian pre-school children | 2009 | Adeniyi et al | Wrong Study Outcome |
| 2 | Assessing Dental Caries and Related Factors in 12‑Year‑Old Nigerian School Children: Report from a Southeastern State | 2020 | Akaji et al | Wrong Study Outcome |
| 3 | Perceived Oral Hygiene Status, Dental Service Utilization and Treatment Needs of Medical and Nursing Students at A Tertiary Institution in Nigeria: a Cross-sectional Study | 2025 | Alade at al | Wrong Study Outcome |
| 4 | Caries Experience Among School Children In Enugu, Nigeria | 2010 | Okoye et al | Incomplete or missing data |
| 5 | Preventive oral health practices of school pupils in Southern Nigeria | 2014 | Folayan et al | Wrong Study Outcome |
| 6 | Oral Hygiene Practices and Utilization of Dental Practices Among Prison Inmates in Bauchi, North East, Nigeria | 2019 | Idowu et al | Wrong Study Outcome |
| 7 | Oral hygiene practices and utilization of oral healthcare services among in-school adolescents in Calabar, Cross River State, Nigeria | 2020 | Ofili et al | Wrong Study Outcome |
| 8 | Oral Hygiene Knowledge and Practices among Rural and Urban Dwellers in parts of South Eastern Nigeria | 2023 | Okorie et al | Wrong Study Outcome |
| 9 | Dental caries and oral health: an ignored health barrier to learning in Nigerian slums (a cross sectional survey) | 2022 | Olatosi et al | Incomplete or missing data |
| 10 | Sociobehavioural risk factors of dental caries among selected adolescents in Ibadan, Nigeria | 2014 | Ajayi et al | Incomplete or missing data |
| 11 | Dental Caries Prevalence, Restorative Needs and Oral Hygiene Status in Adult Population: A Cross‑Sectional Study among Nurses in Jos University Teaching Hospital, Jos, Nigeria | 2021 | Idowu et al | Incomplete or missing data |
| 12 | Dental caries pattern and predisposing oral hygiene related factors in Nigerian preschool children | 2007 | Sowole et al | Incomplete or missing data |
| 13 | Risk factors for rampant caries in children from Southwestern Nigeria | 2012 | Folayan et al | Wrong Study Outcome |
| 14 | Association between mental health, caries experience and gingival health of adolescents in sub‑urban Nigeria | 2021 | Tantawi et al | Wrong Study Outcome |
| 15 | Effect of a school-based oral health education programme on  use of recommended oral self-care for reducing the risk of  caries by children in Nigeria | 2014 | Esan et al | Wrong Study Outcome |
| 16 | Je ̣̀díje ̣̀dí, free sugar consumption and early childhood caries  experience in Ile-Ife, Nigeria: a cultural dimension to dental  caries risk | 2025 | Folayan et al | Wrong Study Outcome |
| 17 | Knowledge of Caries Preventive Measures for Children among Medical and Dental Students in Benin City | 2022 | Ojie et al | Wrong Study Outcome |
| 18 | Factors Influencing Choice Of Oral Hygiene Products By Dental  Patients In A Nigerian Teaching Hospital | 2017 | Opeodu et al | Wrong Study Outcome |
| 19 | Association Between Family Structure and Oral Health in a Group of Nigerian Children | 2025 | Anago et al | Wrong Study Outcome |
| 20 | Burden of oral diseases and dental treatment needs of an urban  population in Port Harcourt, Rivers State, Nigeria | 2014 | Braimoh et al | Wrong Study Outcome |
| 21 | Oral health status, knowledge of dental caries aetiology, and dental clinic attendance: A comparison of secondary school students in the rural and urban areas of Lagos | 2016 | Soroye 2016 | Wrong Study Outcome |
| 22 | Oral hygiene status and practices among rural dwellers | 2013 | Azodo et al | Wrong Study Outcome |
| 23 | Association between Family Level Influences and Caries  Prevention Views and Practices of School Children in a  Sub-Urban Nigerian Community | 2023 | Adeniyi et al | Wrong Study Outcome |
| 24 | Practices regarding oral hygiene among individuals with disabilities attending a special needs institution in Northwestern Nigeria | 2023 | Ogbeide et al | Wrong Study Outcome |
| 25 | Association between early childhood caries and parental educational status among children in Ile-Ife, Nigeria | 2025 | Folayan et al | Secondary data |
| 26 | Associations between the use of caries preventive methods and ECC experience in Ile-Ife, Nigeria: a cross-sectional study | 2025 | Folayan et al | Secondary data |
