## Supplemental Files 1 - 4 for "Association between fluoridated toothpaste use and dental caries in Nigeria: A systematic review and meta-analysis": Supplemetal File 2.docx

**PubMed 30**

("Prevalence"[MeSH Terms:noexp] OR ("epidemiology"[MeSH Subheading] OR "epidemiology"[All Fields] OR "Prevalence"[All Fields] OR "Prevalence"[MeSH Terms] OR "prevalance"[All Fields] OR "prevalences"[All Fields] OR "prevalence s"[All Fields] OR "prevalent"[All Fields] OR "prevalently"[All Fields] OR "prevalents"[All Fields] OR ("epidemiologies"[All Fields] OR "epidemiology"[MeSH Subheading] OR "epidemiology"[All Fields] OR "epidemiology"[MeSH Terms] OR "epidemiology s"[All Fields]) OR ("proportion"[All Fields] OR "proportions"[All Fields]) OR ("epidemiology"[MeSH Subheading] OR "epidemiology"[All Fields] OR "incidence"[All Fields] OR "incidence"[MeSH Terms] OR "incidences"[All Fields] OR "incident"[All Fields] OR "incidents"[All Fields]))) AND ("Fluoride Treatment"[MeSH Terms:noexp] OR ("toothpaste s"[All Fields] OR "toothpastes"[Supplementary Concept] OR "toothpastes"[All Fields] OR "toothpaste"[All Fields] OR "toothpastes"[MeSH Terms] OR ("flouridated"[All Fields] AND ("toothpaste s"[All Fields] OR "toothpastes"[Supplementary Concept] OR "toothpastes"[All Fields] OR "toothpaste"[All Fields] OR "toothpastes"[MeSH Terms])) OR "Dentifrices"[All Fields] OR ("toothpaste s"[All Fields] OR "toothpastes"[Supplementary Concept] OR "toothpastes"[All Fields] OR "toothpaste"[All Fields] OR "toothpastes"[MeSH Terms]))) AND ("Dental Caries"[MeSH Terms:noexp] OR ("carie"[All Fields] OR "Dental Caries"[MeSH Terms] OR ("dental"[All Fields] AND "caries"[All Fields]) OR "Dental Caries"[All Fields] OR "caries"[All Fields] OR "Dental Caries"[All Fields] OR "tooth decay"[All Fields] OR "dental decay"[All Fields] OR "hole"[All Fields] OR "tooth cavity"[All Fields] OR "dental cavities"[All Fields])) AND ("Nigeria"[MeSH Terms:noexp] OR ("Nigeria"[MeSH Terms] OR "Nigeria"[All Fields] OR "nigeria s"[All Fields]))

**Scopus 23**

("Prevalence" OR ("epidemiology" OR "Prevalence" OR "Prevalence" OR "prevalance" OR "prevalences" OR "prevalence s" OR "prevalent" OR "prevalently" OR "prevalents" OR ("epidemiologies" OR "epidemiology" OR "epidemiology" OR "epidemiology" OR "epidemiology s") OR ("proportion" OR "proportions") OR ("epidemiology" OR "epidemiology" OR "incidence" OR "incidence" OR "incidences" OR "incident" OR "incidents"))) AND ("Fluoride Treatment" OR ("toothpaste s" OR "toothpastes" OR "toothpastes" OR "toothpaste" OR "toothpastes" OR ("flouridated" AND ("toothpaste s" OR "toothpastes" OR "toothpastes" OR "toothpaste" OR "toothpastes")) OR "Dentifrices" OR ("toothpaste s" OR "toothpastes" OR "toothpastes" OR "toothpaste" OR "toothpastes"))) AND ("Dental Caries" OR ("carie" OR "Dental Caries" OR ("dental" AND "caries") OR "Dental Caries" OR "caries" OR "Dental Caries" OR "tooth decay" OR "dental decay" OR "hole" OR "tooth cavity" OR "dental cavities")) AND ("Nigeria" OR ("Nigeria" OR "Nigeria" OR "nigeria s"))

**EBSCOHost 56**

("Prevalence" OR ("epidemiology" OR "Prevalence" OR "Prevalence" OR "prevalance" OR "prevalences" OR "prevalence s" OR "prevalent" OR "prevalently" OR "prevalents" OR ("epidemiologies" OR "epidemiology" OR "epidemiology" OR "epidemiology" OR "epidemiology s") OR ("proportion" OR "proportions") OR ("epidemiology" OR "epidemiology" OR "incidence" OR "incidence" OR "incidences" OR "incident" OR "incidents"))) AND ("Fluoride Treatment" OR ("toothpaste s" OR "toothpastes" OR "toothpastes" OR "toothpaste" OR "toothpastes" OR ("flouridated" AND ("toothpaste s" OR "toothpastes" OR "toothpastes" OR "toothpaste" OR "toothpastes")) OR "Dentifrices" OR ("toothpaste s" OR "toothpastes" OR "toothpastes" OR "toothpaste" OR "toothpastes"))) AND ("Dental Caries" OR ("carie" OR "Dental Caries" OR ("dental" AND "caries") OR "Dental Caries" OR "caries" OR "Dental Caries" OR "tooth decay" OR "dental decay" OR "hole" OR "tooth cavity" OR "dental cavities")) AND ("Nigeria" OR ("Nigeria" OR "Nigeria" OR "nigeria s"))

**Web Of Science 20**

("Prevalence" OR ("epidemiology" OR "Prevalence" OR "Prevalence" OR "prevalance" OR "prevalences" OR "prevalence s" OR "prevalent" OR "prevalently" OR "prevalents" OR ("epidemiologies" OR "epidemiology" OR "epidemiology" OR "epidemiology" OR "epidemiology s") OR ("proportion" OR "proportions") OR ("epidemiology" OR "epidemiology" OR "incidence" OR "incidence" OR "incidences" OR "incident" OR "incidents"))) AND ("Fluoride Treatment" OR ("toothpaste s" OR "toothpastes" OR "toothpastes" OR "toothpaste" OR "toothpastes" OR ("flouridated" AND ("toothpaste s" OR "toothpastes" OR "toothpastes" OR "toothpaste" OR "toothpastes")) OR "Dentifrices" OR ("toothpaste s" OR "toothpastes" OR "toothpastes" OR "toothpaste" OR "toothpastes"))) AND ("Dental Caries" OR ("carie" OR "Dental Caries" OR ("dental" AND "caries") OR "Dental Caries" OR "caries" OR "Dental Caries" OR "tooth decay" OR "dental decay" OR "hole" OR "tooth cavity" OR "dental cavities")) AND ("Nigeria" OR ("Nigeria" OR "Nigeria" OR "nigeria s"))

**Embase 25**

("Prevalence" OR ("epidemiology" OR "Prevalence" OR "Prevalence" OR "prevalance" OR "prevalences" OR "prevalence s" OR "prevalent" OR "prevalently" OR "prevalents" OR ("epidemiologies" OR "epidemiology" OR "epidemiology" OR "epidemiology" OR "epidemiology s") OR ("proportion" OR "proportions") OR ("epidemiology" OR "epidemiology" OR "incidence" OR "incidence" OR "incidences" OR "incident" OR "incidents"))) AND ("Fluoride Treatment" OR ("toothpaste s" OR "toothpastes" OR "toothpastes" OR "toothpaste" OR "toothpastes" OR ("flouridated" AND ("toothpaste s" OR "toothpastes" OR "toothpastes" OR "toothpaste" OR "toothpastes")) OR "Dentifrices" OR ("toothpaste s" OR "toothpastes" OR "toothpastes" OR "toothpaste" OR "toothpastes"))) AND ("Dental Caries" OR ("carie" OR "Dental Caries" OR ("dental" AND "caries") OR "Dental Caries" OR "caries" OR "Dental Caries" OR "tooth decay" OR "dental decay" OR "hole" OR "tooth cavity" OR "dental cavities")) AND ("Nigeria" OR ("Nigeria" OR "Nigeria" OR "nigeria s"))

**African Journals Online 23**

Caries AND toothpaste

**Google Scholar	999**

Caries AND toothpaste  AND Nigeria

**West Africa College of Surgeons 8**

Caries AND toothpaste
